# Digitizing ECG image: new fully automated method and open-source software code

**DOI:** 10.1101/2021.07.13.21260461

**Authors:** Julian D. Fortune, Natalie E. Coppa, Kazi T. Haq, Hetal Patel, Larisa G. Tereshchenko

## Abstract

**Background:** We aimed to develop and validate an automated, open-source code ECG-digitizing tool and assess agreements of ECG measurements across three types of median beats, comprised of digitally recorded, simultaneous and asynchronous ECG leads and digitized asynchronous ECG leads.

**Methods:** We used the data of clinical studies participants (n=230; mean age 30±15 y; 25% female; 52% had the cardiovascular disease) with available both digitally recorded and printed on paper and then scanned ECGs, split into development (n=150) and validation (n=80) datasets. The agreement between ECG and VCG measurements on the digitally recorded time-coherent median beat, representative asynchronous digitized, and digitally recorded beats was assessed by Bland-Altman analysis.

**Results:** Agreement between digitally recorded and digitized representative beat was high [area spatial ventricular gradient (SVG) elevation bias 2.5(95% limits of agreement [LOA] -7.9-13.0)°; precision 96.8%; inter-class correlation [ICC] 0.988; Lin’s concordance coefficient ρ_c_ 0.97(95% confidence interval [CI] 0.95-0.98)]. Agreement between digitally recorded asynchronous and time-coherent median beats was moderate for area-based VCG metrics (spatial QRS-T angle bias 1.4(95%LOA -33.2-30.3)°; precision 94.8%; ICC 0.95; Lin’s concordance coefficient ρ_c_ 0.90(95%CI 0.82-0.95)], but poor for peak-based VCG metrics of global electrical heterogeneity.

**Conclusions:** We developed and validated an open-source software tool for paper-ECG digitization. Asynchronous ECG leads are the primary source of disagreement in measurements on digitally recorded and digitized ECGs.

## 1. Introduction

An electrocardiogram (ECG) is a ubiquitous, inexpensive, noninvasive diagnostic tool. ECG is widely used in everyday clinical practice around the globe. Besides clinical practice, ECG is a common phenotype used in “big data” genomics[1] and artificial intelligence (AI) studies.[2] ECG characterizes the electrophysiological substrate of cardiovascular diseases and can be easily recorded in hundreds of thousands of people, delivering the required statistical power for analyses. Rare clinical outcomes (e.g., sudden cardiac death) require a large sample size and long-term follow-up.[3] In modern days, ECG is routinely saved and stored as a digital file. However, until the end of the 20^th^ century, ECG was recorded on paper. Many large-scale clinical studies that were conducted in the 20^th^ century collected thousands of unique paper-printed ECGs. Paper-printed ECGs have to be scanned and digitized to make them available for future AI studies[4] and the application of novel analytical approaches, such as global electrical heterogeneity (GEH) measurement.[5]

Several ECG-digitizing tools have been previously developed.[6] The most widely used is the ECGScan, a patented, commercially available computer application.[7] The ECGScan utilized the concept of active contours and dynamic programming, providing the possibility for the user to set anchor points on the ECG waveform’s image manually. While the users usually appreciated the possibility to provide their input, manual “correction” of the ECG waveform was time-consuming and reduced the reproducibility of ECG digitization.[7, 8] Notably, previously, there was no open-source code ECG-digitizing tool available.

It is well-recognized that digitized ECG is inferior to digitally recorded ECG.[6-8] Compared to digitally recorded ECG, the digitized ECG inevitably loses the information and should be used only if digitally recorded ECG is not available.[9] Of note, historic ECG machines and modern standard 3×4 display of a 12-lead ECG printout show different ECG leads in different moments of time. Modern standards require ECG measurements performed on a median beat created out of simultaneously recorded ECG leads.[10] Such requirements cannot be fulfilled using asynchronous digitized ECG leads. However, the degree of agreement between ECG measurements performed on a median beat comprised of simultaneously versus asynchronously recorded ECG leads has not been assessed.

Recent advancements in AI algorithms opened avenues for further development of ECG-digitizing tools. In the present study, we aimed to develop and validate an automated, open-source code ECG-digitizing tool and assess agreements of standard ECG and vectorcardiographic (VCG) GEH measurements across three types of median beats, comprised of (1) digitally recorded, simultaneous ECG leads, (2) digitally recorded, asynchronous ECG leads, and (3) digitized, asynchronous ECG leads.

## 2. Methods

### 2.1. Study population and ECG recording

For the development and validation of the algorithm, we used the data of clinical studies conducted at the Oregon Health & Science University (OHSU), with available both (1) digitally recorded ECGs and (2) printed on paper ECGs.[11-14] All studies were approved by the OHSU Institutional Review Board, and all participants signed informed consent before entering the study.

Routine resting 10-second 12-lead ECGs were recorded using a MAC 5500 HD ECG system (General Electric (GE) Healthcare Technologies, Marquette Electronics, Milwaukee, WI). ECGs were printed with a standard output mode: at the paper speed of 25 mm/s and 10 mm/mV calibration, in a 3×4 display mode. Consecutive 2.5 seconds were displayed on a printed ECG record so that the first 2.5 seconds displayed leads I, II, III, the second 2.5 seconds displayed leads aVR, aVL, aVF, the third 2.5 seconds displayed leads V1, V2, V3, and the final 2.5 seconds displayed leads V4, V5, and V6.

In the present study, we excluded participants with atrial fibrillation of flutter recorded on a 12-lead ECG.

Paper ECG printouts were scanned at 600 dpi resolution, color mode.

We used the Magellan ECG Research Workstation V2 (GE Marquette Electronics, Milwaukee, WI) to obtain the raw digital ECG signal with the sampling rate of 500 Hz and 1 µV amplitude resolution.

### 2.2. Algorithm development and description

The algorithm and open-source software code, written in Python (Python Software Foundation, Delaware, US), are provided at https://github.com/Tereshchenkolab/paper-ecg. Figure 1 shows the flowchart of the developed algorithm. The complex task of digitizing a scan containing many ECG leads was subdivided into 3 steps to clarify the scope (Table 1).

**Table 1.** Input and output in each step of the complex task of digitizing an ECG scan.

| Process steps | Input | Output |
| --- | --- | --- |
| Digitization | Color image | Scalar or tensor data |
| Detection | Color image | Binary image |
| Extraction | Binary image | Scalar or tensor data |

**Figure 1.**
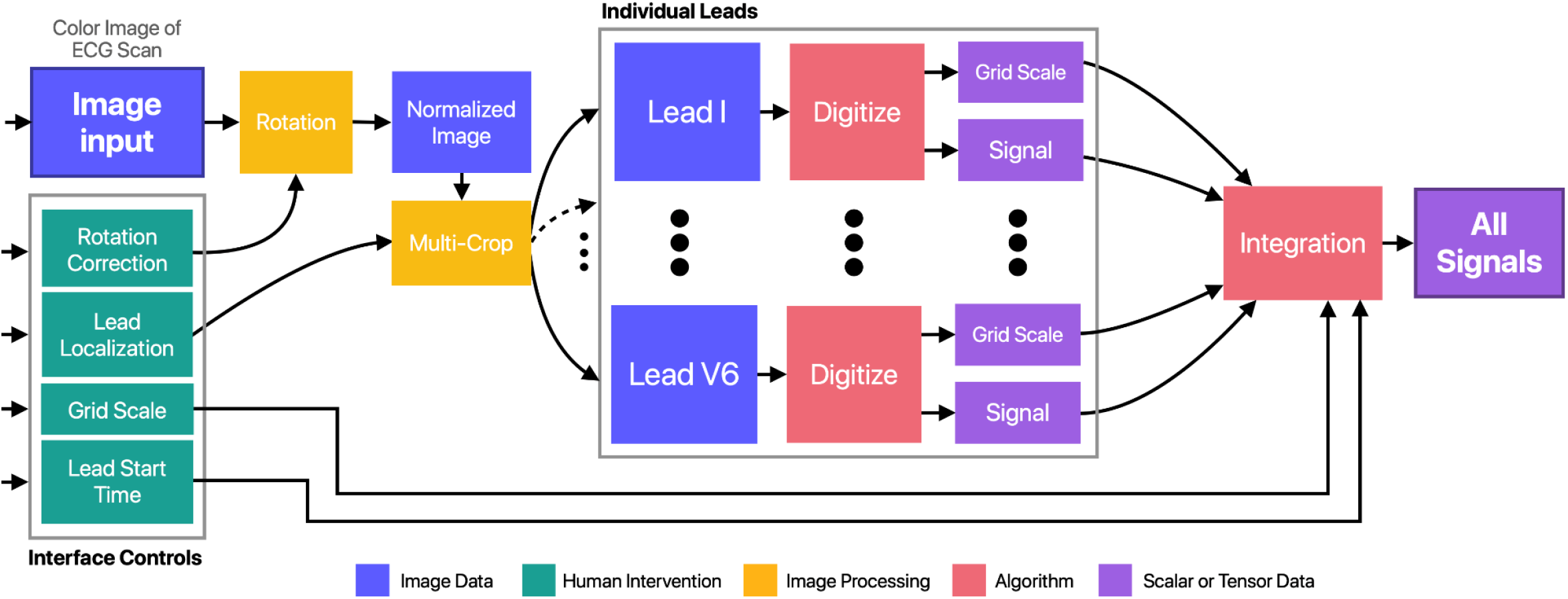
Flowchart of the developed algorithm.

The digitization process involves two parallel processes, grid digitization and signal digitization (Figures 2 and 3). The detection involves converting a color image of a cropped ECG lead to a binary image such that only pixels of interest are *True*. The extraction takes the binary image as an input and generates the relevant output: a scalar estimate of the spacing for the grid and an array of height values for the signal.

**Figure 2.**
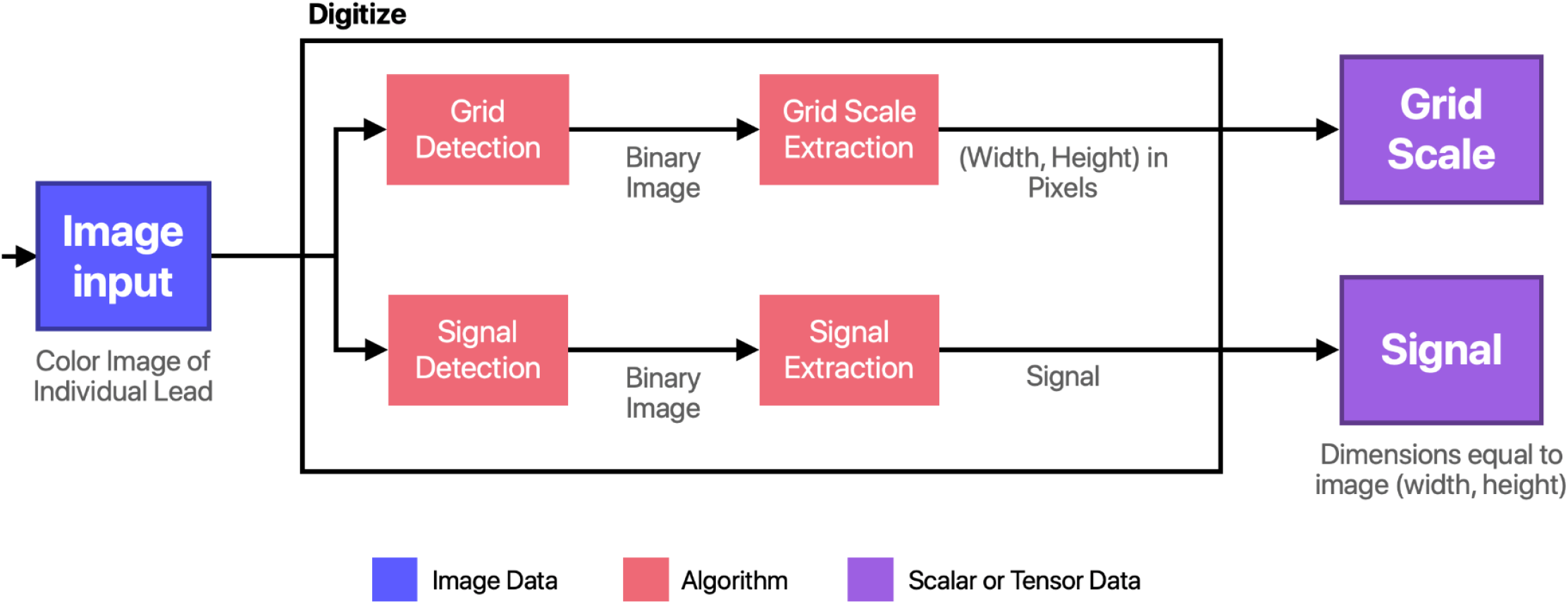
Flowchart of the Digitization Module.

**Figure 3.**
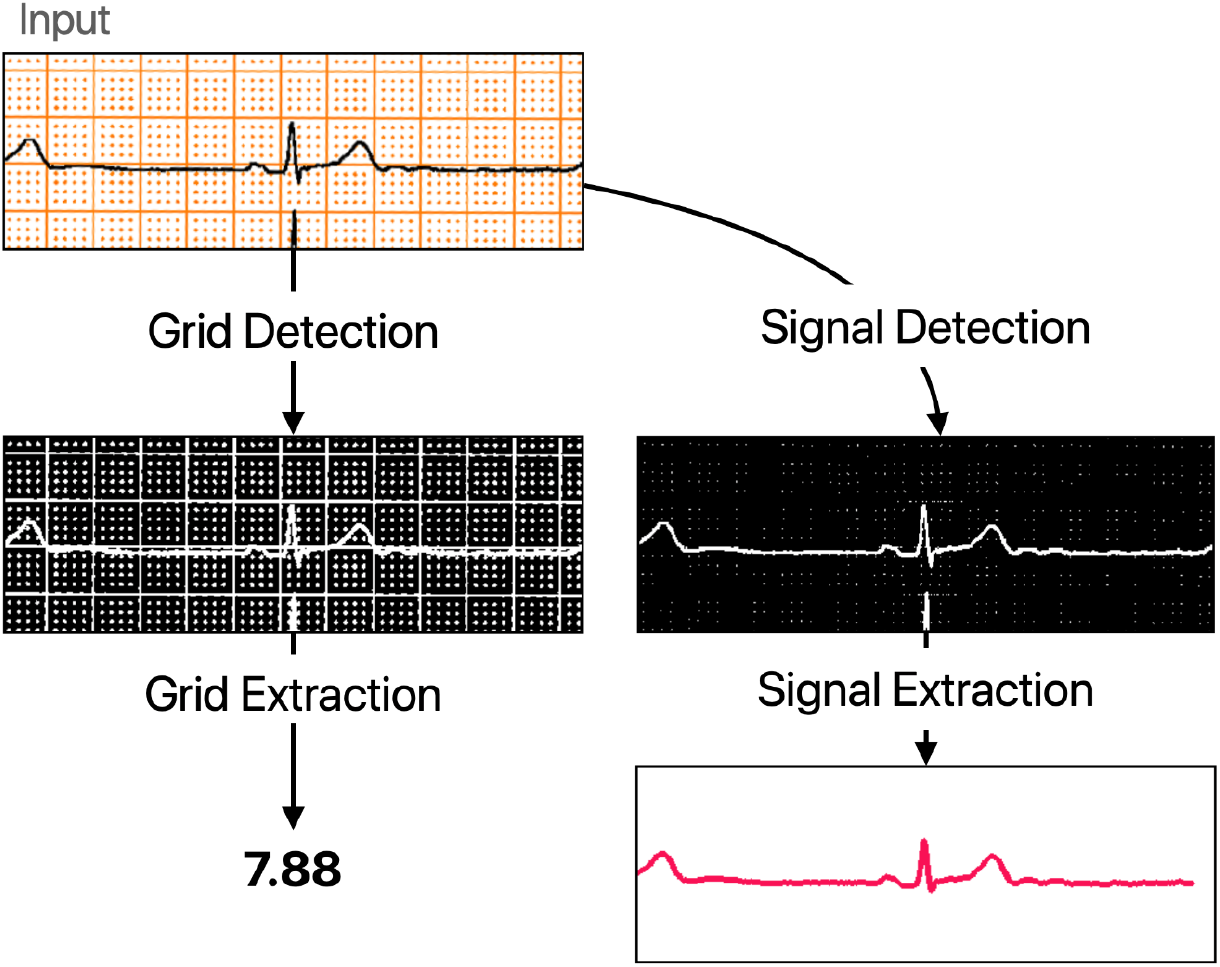
The overview and the representative example of the digitization process.

#### 2.2.1. Image preparation and digitization

First, the application prepares the ECG scan, allows rotation/normalization, crops individual ECG leads, and integrates the digitized data from all leads (Figure 4). The inputs consist of the color image of the scanned ECG and user inputs for rotation, lead locations, lead start time, and grid scale. The input image is rotated and cropped based on the user inputs to produce between one and twelve images of individual leads. Each of these lead images goes through the same digitization process, and the digitization outputs are all joined together into a matrix which is output to a file.

**Figure 4.**
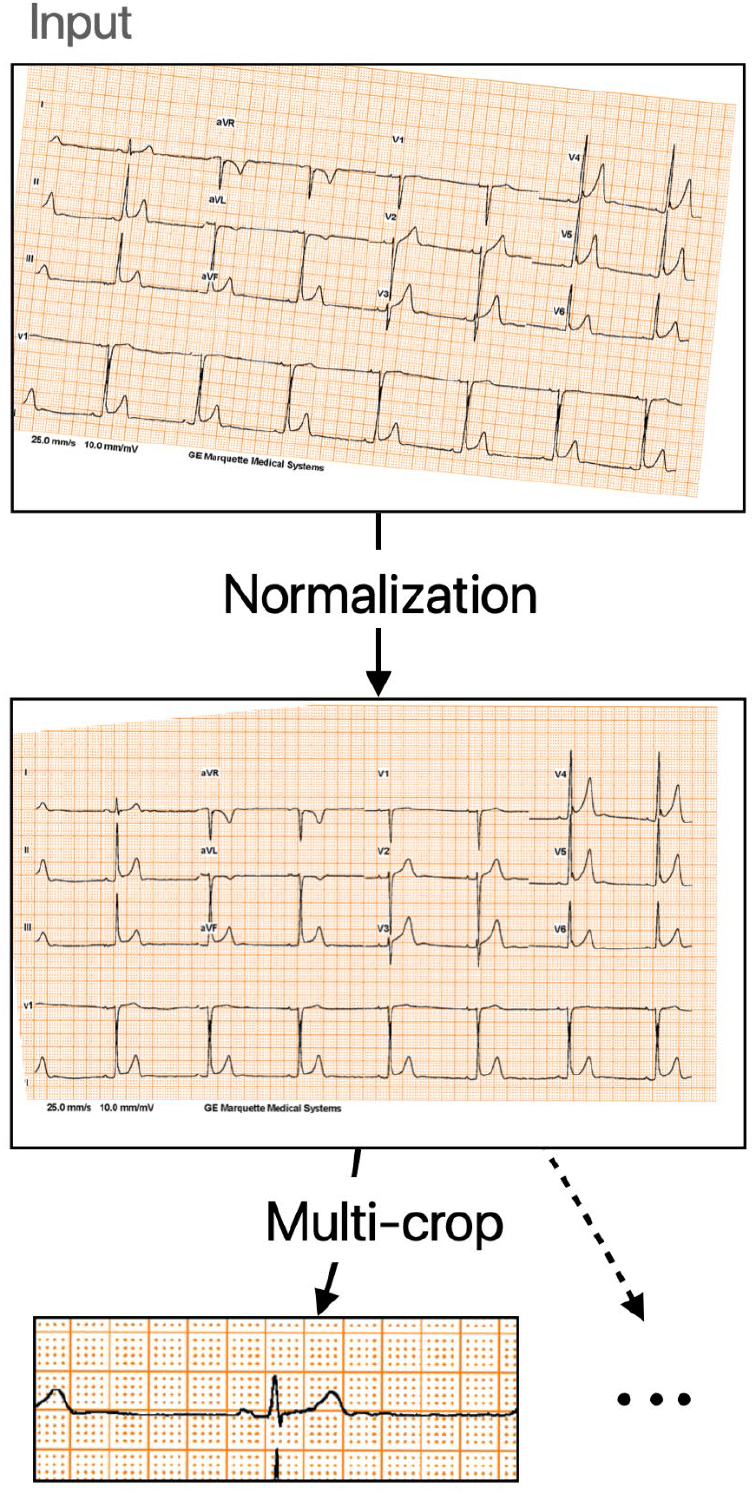
Preparation of the ECG mage for digitizing. The input image is rotated by 5.5 degrees (specified by user or autorotation) to produce the normalized image. Then, the normalized image is cropped to x=73, y=237, width=488, and height=165 to produce the cropped lead image. The application may produce up to 12 cropped lead images, but only one is shown for brevity.

A Graphical User Interface (GUI) provides controls for users to perform rotation correction, input the grid scale, localize leads, and input lead start times. It also offers visual feedback by overlaying the extracted signals on the original images after digitization is complete.

The GUI was developed using PyQt5, a set of Python bindings for the Qt framework.[15] The design goal was to keep the GUI simple and require as little human interaction as possible. The GUI consists of three main components: the menu bar, image view, and control panel (Figure 5). Figure 6 depicts the typical user workflow when using the application.

**Figure 5:**
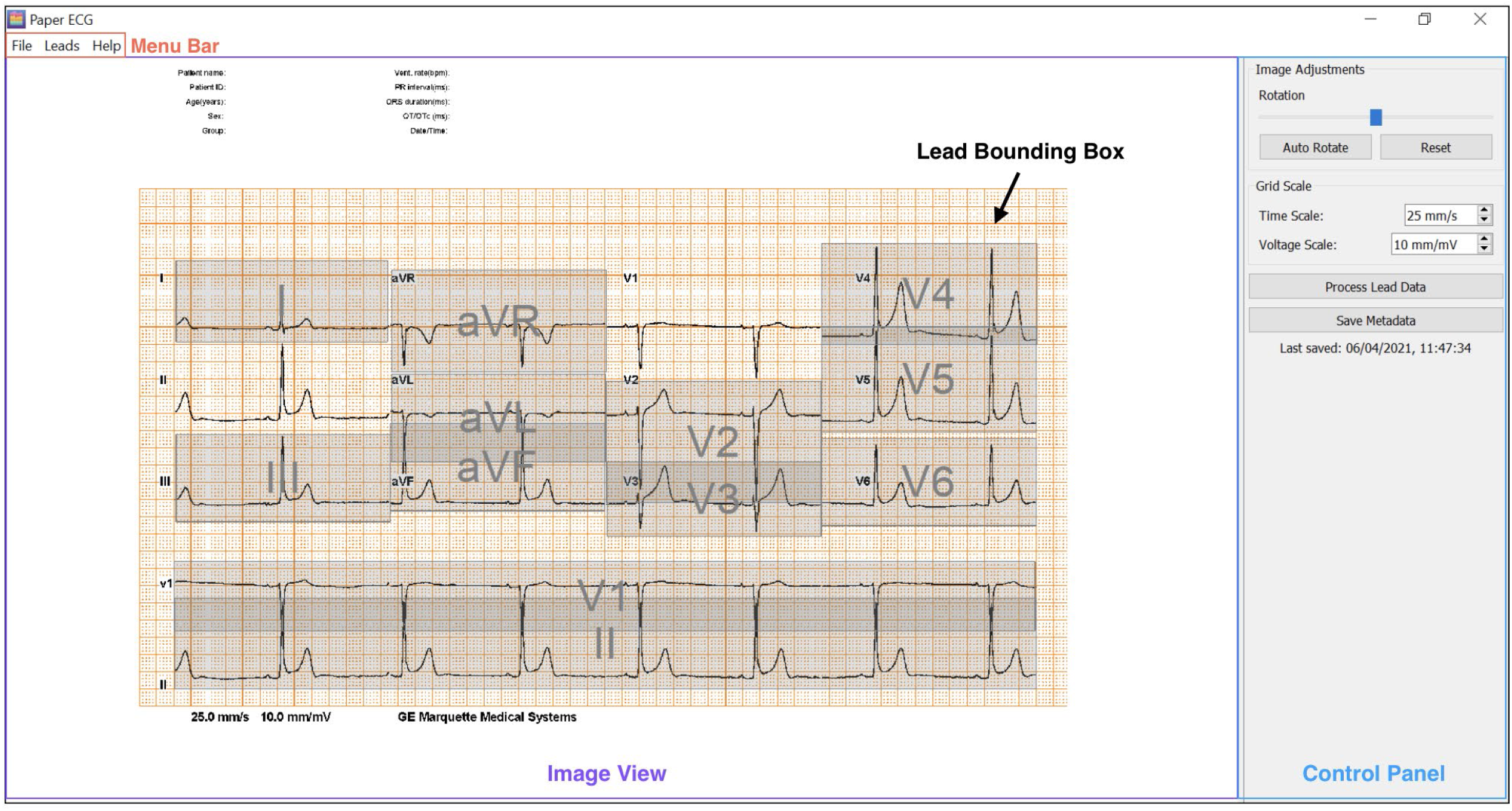
The Graphical User Interface (GUI). The menu bar allows users to open and close image files, add lead bounding boxes, and find help using the application. The image view displays the image opened by the user. This is where users add bounding boxes to perform lead localization. The control panel houses the main controls for the application. The “global” view, which is displayed when no lead bounding boxes are selected, has controls for rotating the image, entering the grid scales, performing digitization, and saving the user’s current annotations. When a lead bounding box is selected, the control panel switches to the “lead” view, where users can enter the lead’s start time or delete the lead’s bounding box.

**Figure 6.**
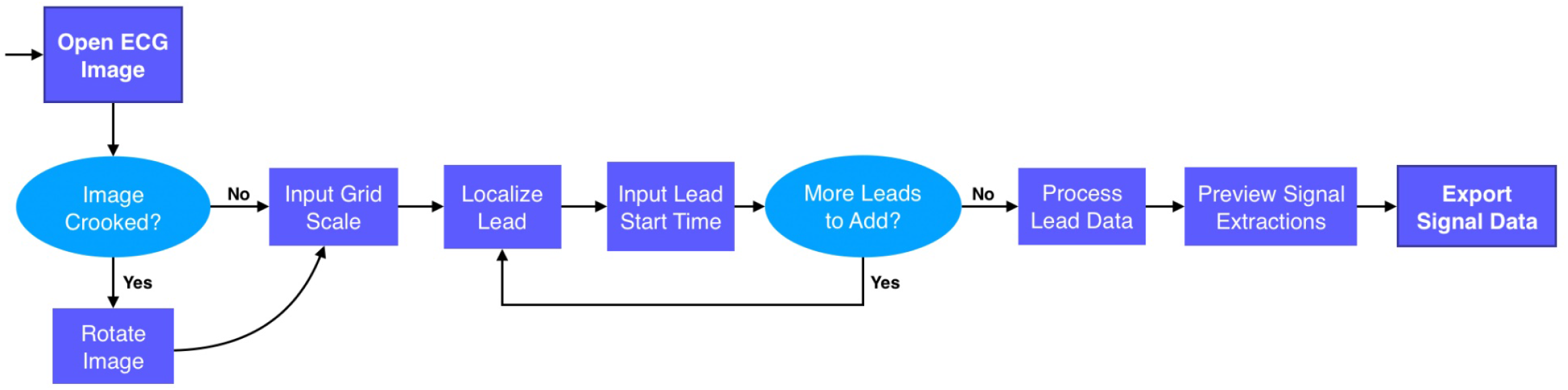
The typical user workflow when using the application.

Users can adjust the image rotation using the slider in the application’s control panel. The rotation is implemented via OpenCV [16] and maintains the original size while filling any empty pixels with white.

The user inputs include bounding boxes associated with the standard twelve leads. These bounding boxes are applied to the image after rotation to create color images of single leads that are inputs to the digitization process (Figure 7A).

**Figure 7:**
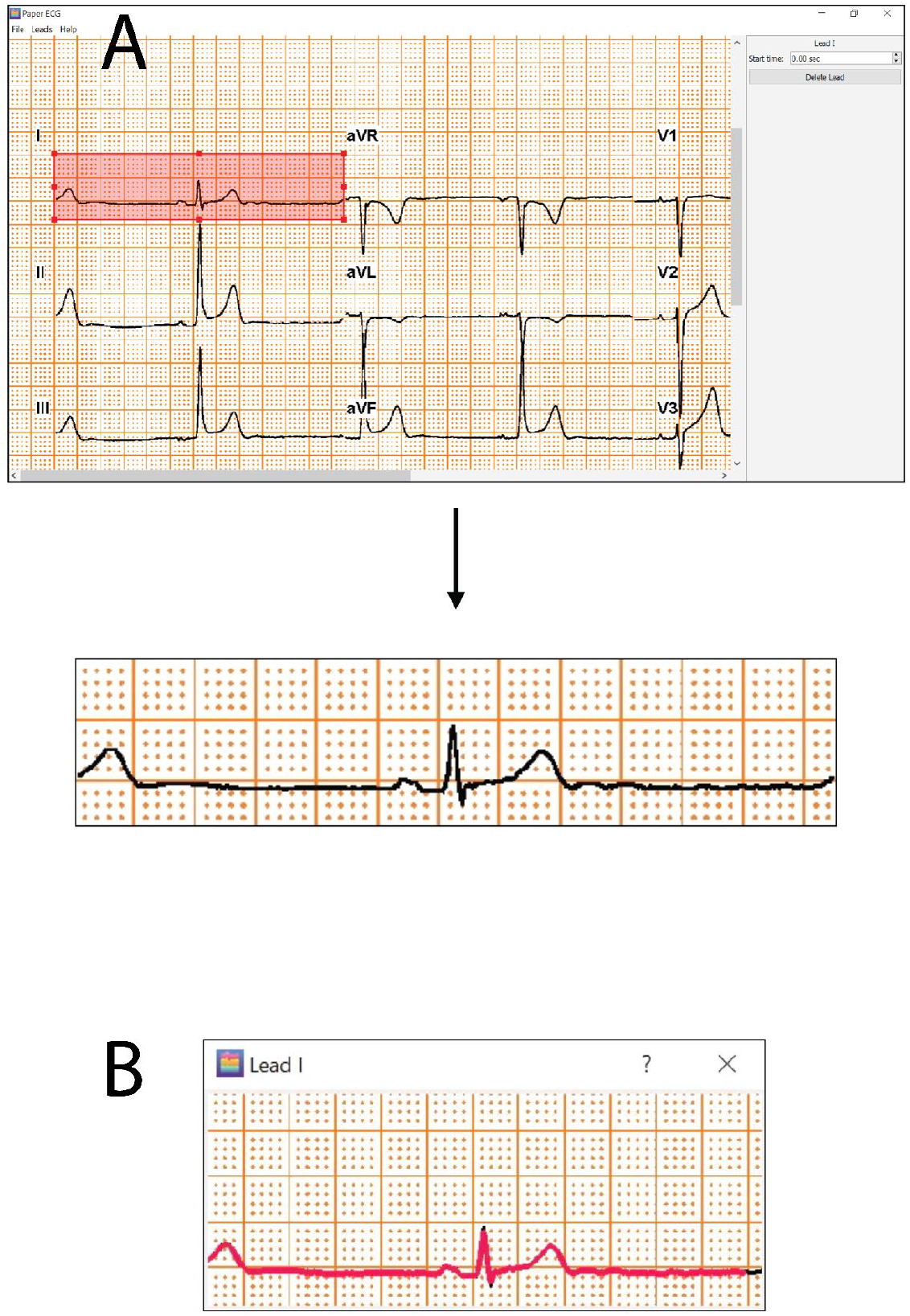
Representative example of (A). Lead cropping. (B). The preview images for each lead, showing the extracted digital signal (red) drawn over the top of the original ECG image signal (black).

Next, the cropped lead images are passed to the digitization module, after which there is an integration process that involves post-processing, aligning, and padding each lead. The integration process takes the un-altered signal (i.e., pixel height locations for every column in the image) and grid sizing from the digitization module for each lead the user has elected to digitize and creates a matrix with a column for each lead to output to a file.

The grid spacings from all leads where grid digitization was successful are averaged to get a more robust estimate and provide an estimate in cases where grid digitization failed for a lead image. Then, the same post-processing is applied to the outputs from the digitization process for all leads.

The first step of post-processing is to zero the signal since the raw signal is based on an origin located at the top left corner of the image. The zero-ed signal is produced by subtracting the median of the signal from each point in the signal.

The second step of post-processing is to scale the signal. The scaling factor, shown in equation 1, is the product of: the inverse of the grid size, which converts the signal from units of pixels to millimeters; the inverse of the user-supplied voltage scale, which converts units of millimeters to millivolts; and 1000 to convert from millivolts to microvolts.

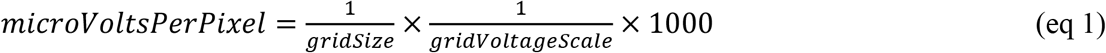

After post-processing each signal, the signals are padded with zeros such that every signal has the same length. First, the user-supplied start times are used to pad signals on the left side. The number of pixels to pad is determined by dividing the start time (in seconds) by the horizontal scaling factor, defined in equation 2.

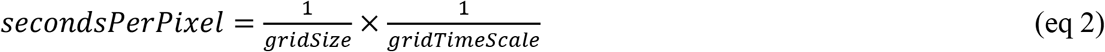

Finally, all signals are padded with zeroes on the right such that all are the same length.

After integration, the signal data is passed back to the GUI for the user to preview (Figure 7B) and save to a file.

#### 2.2.2 Grid detection

The robust nature of the grid extraction method allows for a simplistic, and therefore efficient, threshold process: the image is converted to grayscale, the white point is normalized using the median intensity, and a static threshold is used to generate a binary image. The grayscale conversion is performed by the OpenCV implementation *(cvtColor())*, which calculates the intensity, or “gray” value, as shown in equation 3.[17]

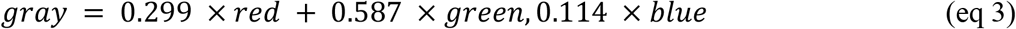

Next, the histogram of the grayscale image is computed, and the median of the histogram is assumed to be a representative value for white. Then, the pixels’ values are multiplied (element-wise) by the median value of the histogram divided by 255 to create a white-point-adjusted image. Lastly, all pixels with intensities above 230 are selected as *True* in the binary image. This value, 230, was chosen based on manual tuning on a subset of cropped ECG lead images from various scans used to verify the code was performing as expected (referred to henceforth as the “Development dataset”).

#### 2.2.3. Grid extraction

The grid extraction is performed in both horizontal and vertical directions separately by summing the *True* pixels in a row or column across the whole image, then estimating the fundamental frequency of this signal (Figure 8).

**Figure 8.**
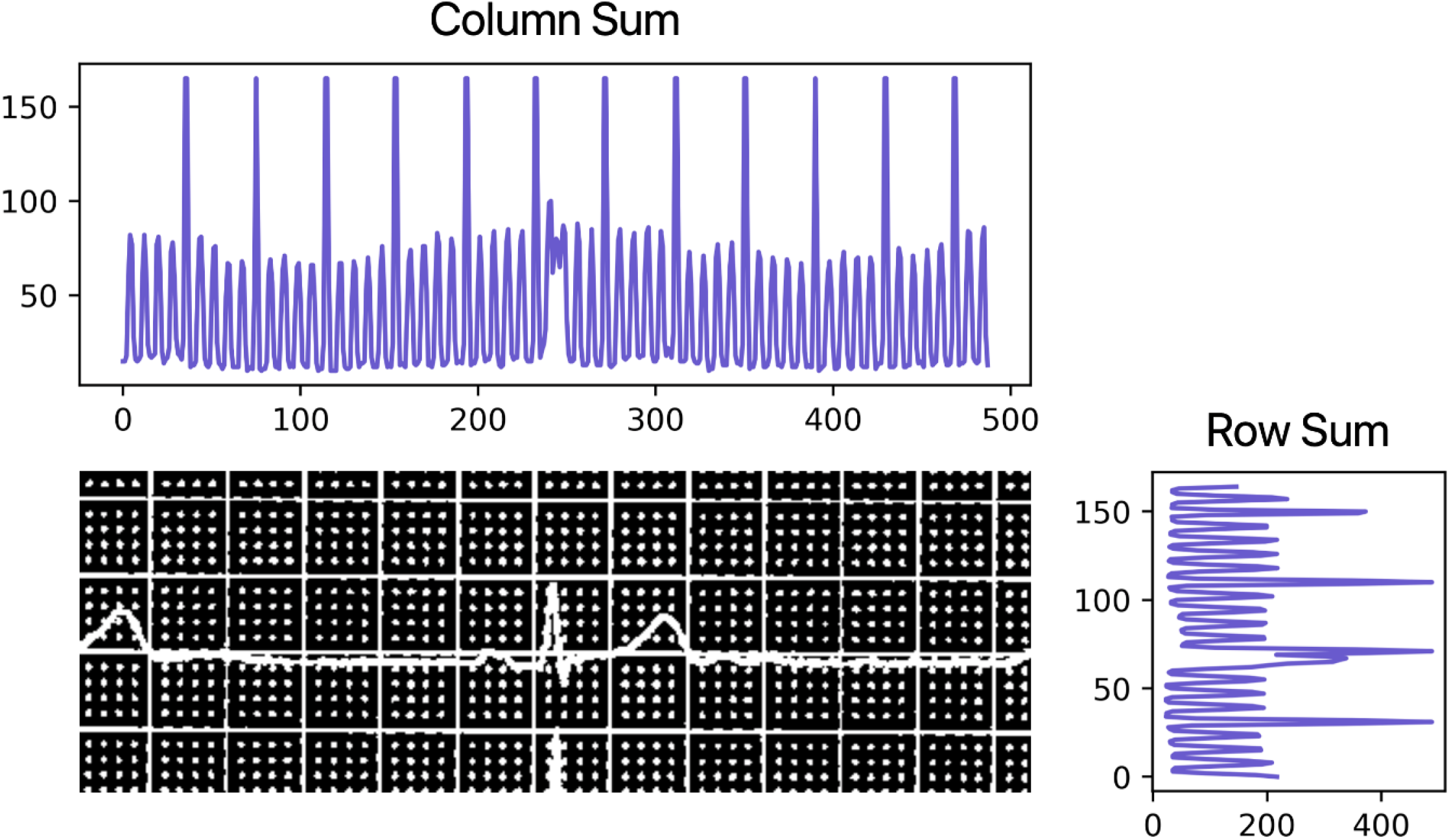
Grid extraction. The column count (a) and the row count (c) for the binary image input (b) are shown. The count signals have a comb-like shape where teeth occur at grid line locations. In this example, the grid involves some dotted lines, which results in lower counts for those rows or columns, but the results of frequency analysis are not different from a grid with all solid lines.

The fundamental period (the period of the fundamental frequency) of the column and row count signals is obtained via computing the autocorrelation of the signal and selecting the first peak in the autocorrelation signal with a height greater than 0.3 (the range of autocorrelation is [-1, 1]), and prominence of 0.05, implemented via the SciPy library.[18] These parameters were chosen via manual tuning using the Development dataset.

A step size of one pixel is used for autocorrelation, and this may result in a large error when the grid size is on the order of one or ten pixels. Therefore, a quadratic function is fitted to the three points around and including the peak, and the maximum over that function (with a precision of 0.01 pixels) is taken as the true period of the grid.

The pixel-counting approach to grid extraction is robust to slight changes in rotation but may suffer when the rotation is sufficiently erroneous. When multiple horizontal grid lines are counted in the same row of the image, or multiple vertical grid lines are counted in the same column of the image, the signal will not have sharp peaks, which can cause fundamental period analysis to fail in finding an acceptable peak. The row analysis is more sensitive because it involves longer lines, which increases the likelihood of overlap and, in turn, diminishes peaks in the count signal. Therefore, the output from grid extraction is the period estimate from the columns when available, and the period estimate from the rows only when the column estimation fails. It is also possible for extraction to fail entirely, which may indicate a lack of grid lines, an erroneous rotation of the image that prevents lines from being distinguished from one another, or a highly noisy image.

#### 2.2.4. Signal detection

The signal detection relies upon the grid extraction module to detect when a grid is or is not present in order to adaptively set the threshold. The process begins by converting the image to grayscale using equation 1, selecting a starting threshold based on Otsu’s method,[19] and creating a binary image based on that threshold. Next, iteratively, grid extraction is applied and, if grid extractions succeed (i.e., an estimate for the spacing is returned), this threshold is reduced by 5%, and a new binary image is created. The iteration stops when the reduced threshold is below 60% of the original threshold or grid extraction fails (i.e., the grid is not sufficiently salient in the binary image). The stopping point of 60% was chosen based on the Development dataset. The step size of 5% was chosen to balance run-time with sufficient granularity, again verified on the Development dataset. The final binary image created during the linear search is returned.

#### 2.2.5. Signal Extraction

The signal extraction algorithm is based on the Viterbi dynamic programming (DP) algorithm,[20] where contiguous regions in each column of the image are treated as nodes, and a custom score (cost) function is provided to incentivize paths that are “signal-like.” The first step involves examining each column, or horizontal slice, of the input binary image and marking the center of each region of contiguous *True* pixels. This produces a list of lists of points in each column that are sorted topologically when scanning across the image from left to right.

Next, the DP table is built. First, all the points in the first column of the image are stored in the DP table as base cases with costs of zero. Then, the points in each column are processed, starting with the second column from the left and going to the rightmost column. At each point, possible connections can be made to candidate points in the nearest prior column containing points. This is typically the immediately-prior column, but columns can be empty. Each possible connection has a total cost consisting of the cost to get to the candidate point plus the cost to transition from the candidate point to the current point. The cost function is defined in equation 4,

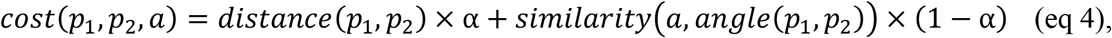

where: p_1_ is the candidate point, p_2_ is the current point, *a* is the angle of the connection arriving at the candidate point (i.e., the instantaneous direction of the path at p_1_), *distance* returns the Euclidean distance between two points, *alpha* is a parameter to control the relative weight of distance and angle alignment, the *similarity* is defined in equation 5, and *angle* is the angle between two points in degrees.

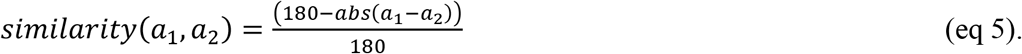

The candidate point with the minimal total cost is selected as the best path to the current point, and this is recorded in the DP table. If no possible connections can be found, the point becomes a base case with a cost of zero.

The cost function is designed to grant a lower cost for points that are closer in the image and penalize points that are distant because the shape of the signal is generally continuous (i.e., does not involve abrupt jumps up or down from one instant to the next) and does not involve large horizontal gaps. The costs function also penalizes large changes in direction by comparing the change in direction from one point to the next because the line is generally relatively smooth. The value of *alpha* 0.5 is selected, as determined by manual examination on the Development dataset.

An example of a DP table is shown in Figure 9, where blue represents a low total cost and maroon a high total cost. Once the DP table has been constructed for all points in the image, a single best path is chosen by searching for the path with the lowest cost in the last 20 columns in the image. Then, backtracking is performed by following the back-references in the entry for each point in the DP table to construct the entire path. There is a final step to convert the list of points to an array of the y values, and where *NaN* values are inserted for any columns where a point is absent, and this array is returned.

**Figure 9.**
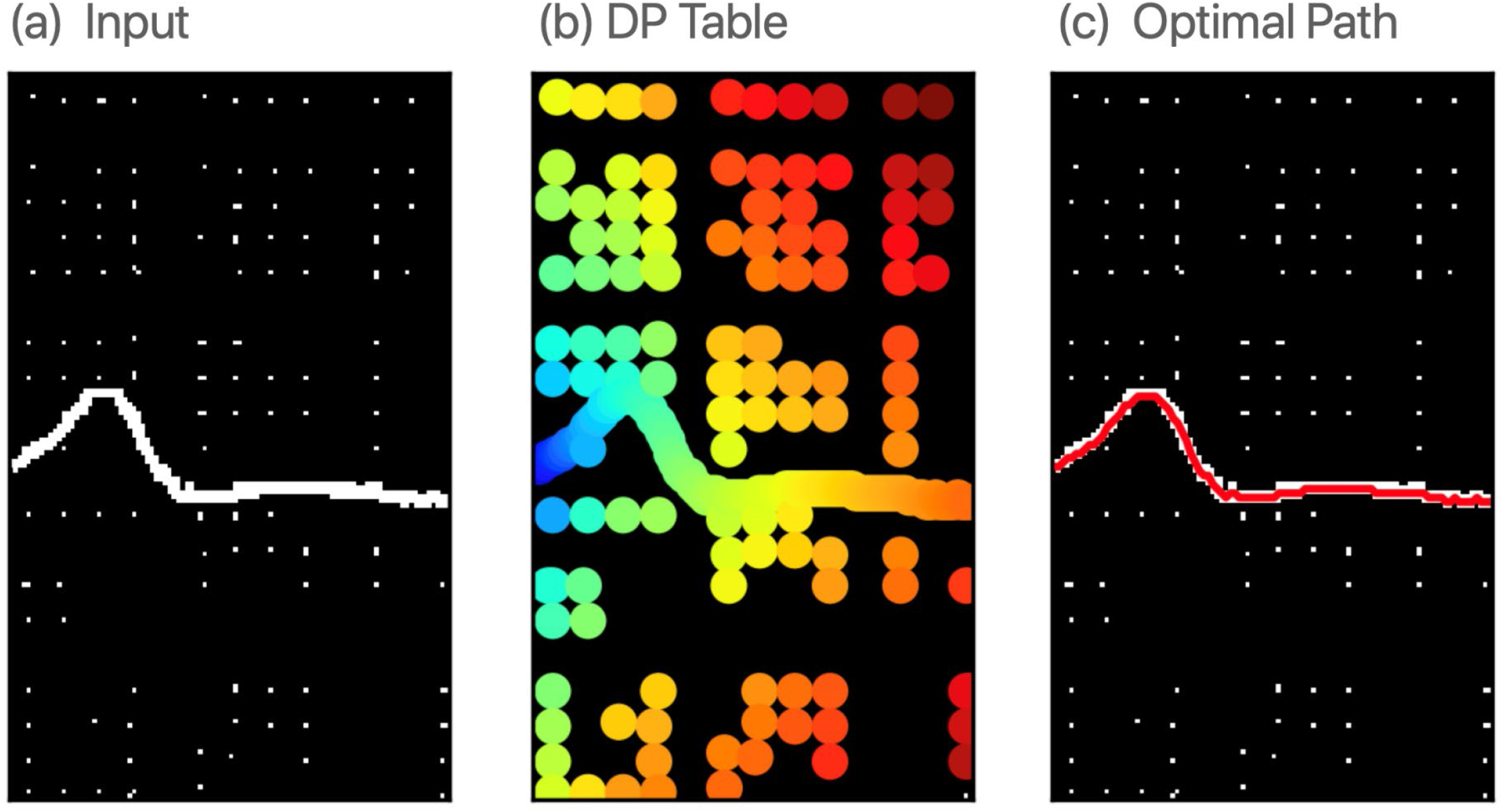
An example of the Viterbi dynamic programming table. The input image (a) is processed by the Viterbi dynamic programming algorithm. The total cost for the path to each point is based on the dynamic programming table (b). The minimal total cost is selected as the best path (c).

### 2.3. Validation of the automated digitizing algorithm

For validation, we assessed the agreement in the standard ECG and VCG GEH measurements, performed on three types of ECG signal: (1) digitally recorded, simultaneous ECG leads, (2) digitally recorded, asynchronous ECG leads, and (3) digitized, asynchronous ECG leads.

#### 2.3.1. ECG and VCG measurements on digitally recorded, simultaneous ECG leads

Using a 10-second, simultaneously recorded 12-lead digital ECG signal, we detected the origin of the heart vector and comprised the time-coherent global XYZ median beat as previously described.[21] Using previously validated algorithms [22, 23], we detected ECG fiducial points on the median beat vector magnitude signal. The accuracy of automated fiducial point detection was confirmed by two investigators (KTH, HP) using visual aid. Next, we measured traditional ECG metrics (PR, QRS, QT, and Bazett-corrected QTc intervals). In addition, VCG GEH metrics (area-based and peak-based spatial ventricular gradient [SVG] magnitude, azimuth, and elevation, spatial QRS-T angle, sum absolute QRST integral [SAIQRST], and vector magnitude QT integral [VMQTi]) were measured as previously described.[21, 24]

#### 2.3.2. ECG and VCG measurements on digitized asynchronous ECG leads

On the digitized ECG signal, we selected one clean representative cardiac beat on each out of eight leads: I, II, V1-V6. Next, the time point of the maximum absolute |dV/dt| QRS value was automatically detected on each selected cardiac beat. All eight cardiac beats were aligned using the detected maximum absolute |dV/dt| time point and converted to XYZ using Kors transformation matrix,[25] which led to the construction of a representative global XYZ beat. Then, the origin of the heart vector was identified.[21] After that, ECG and VCG measurements were performed as described in sub-section 2.3.1.

#### 2.3.3. ECG and VCG measurements on digitally recorded asynchronous ECG leads

To validate the digitization process and to distinguish an error introduced by an asynchronous cardiac beat from the digitization error, we constructed the representative global XYZ beat using the digitally recorded asynchronous ECG signal. We selected the same cardiac beats on the same ECG leads that were used for the analysis of digitized ECG signal. Then, all the signal processing steps described in subsection 2.3.2 were performed as described above.

#### 2.3.4. Assessment of the agreement

The agreement between ECG and VCG measurements on the digitally recorded time-coherent median beat, representative asynchronous digitized beat, and the representative asynchronous digitally recorded beat was assessed by Bland-Altman analysis.[26, 27] We calculated the degree of the agreement as to the bias (the absolute mean difference) with 95% limits of agreement (LOA). The Bradley-Blackwood procedure was used to compare the means and variances of the 2 compared measurements simultaneously.[28] Bonferroni-adjusted significance of the Bradley-Blackwood F-test (*P*<0.001) indicated a statistically significant correlation of compared means and variances, implying biased estimates of the Bland-Altman analysis. Precision was defined as 100% minus relative % bias. Interclass correlation coefficient (ICC), equivalent to Cronbach’s alpha statistic,[29] was calculated for standardized variables (in the scale to mean 0 and variance 1). Lin’s concordance correlation coefficient ρ_c_ was calculated to describe the strength of agreement: >0.99 indicated almost perfect agreement; 0.95–0.99, substantial agreement; 0.90–0.95, moderate agreement; <0.90, poor agreement.

As the SVG azimuth variables are circular (ranging from -180° to +180°), to calculate relative bias and ICC, SVG azimuth variables were transformed by doubling their value and adding 360. The agreement assessment was performed using STATA MP 17.0 (StataCorp LLC, College Station, TX, USA).

## 3. Results

### 3.1. Study population

The study population included 240 subjects. The study dataset was randomly divided into development (n=150; 65%) and validation (n=80; 35%) subsets. There were no differences in clinical and demographic characteristics of subjects included in development and validation datasets (Table 2). Approximately one-quarter of subjects were female, and more than 80% were white. About half of the participants were healthy, and their ECGs were normal, whereas another half had a history of cardiovascular disease and abnormal ECG.

**Table 2.** Study population characteristics.

|  | Development (n=150) | Validation (n=80) |
| --- | --- | --- |
| Age(mean±SD), yrs | 29.7±15.0 | 29.5±13.9 |
| Female, n(%) | 40(26.7) | 17(21.3) |
| White, % | 134(89.3) | 69(86.3) |
| Body mass index(mean±SD), kg/m<sup>2</sup> | 28.8±7.8 | 29.2±8.2 |
| Cardiovascular disease, n(%) | 80(53.3) | 40(50.0) |

### 3.2. Validation results

Examples of a digitized 12-lead ECG are shown in Figures 10 and 11. For further quantitative assessment of the agreement, we excluded ECGs (n=40) with obvious visible errors of the digitization, assessed by two investigators (KTH, HP).

**Figure 10.**
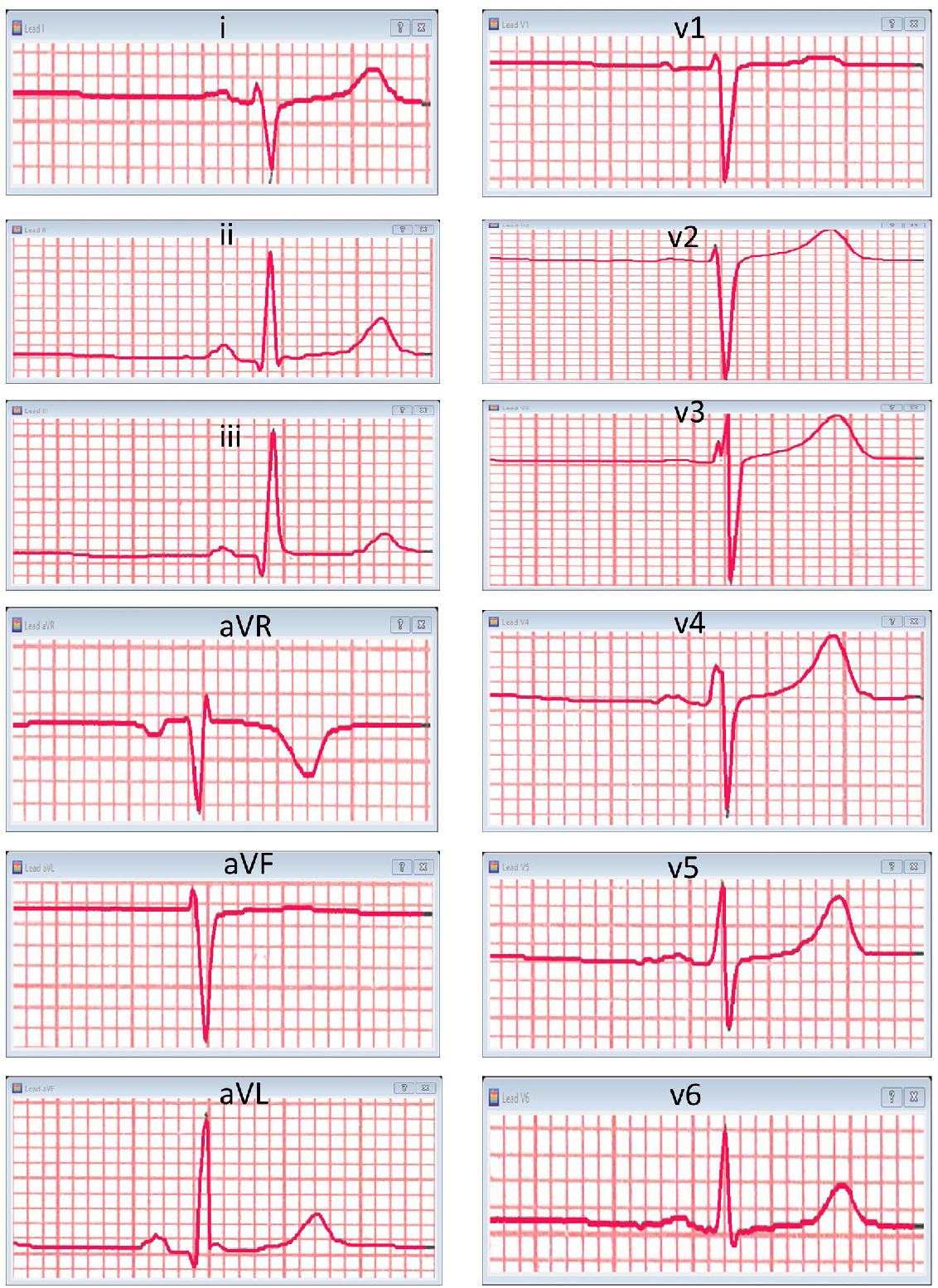
Representative example of digitized 12-lead ECG signal (red) overlapping ECG image waveform (black).

**Figure 11.**
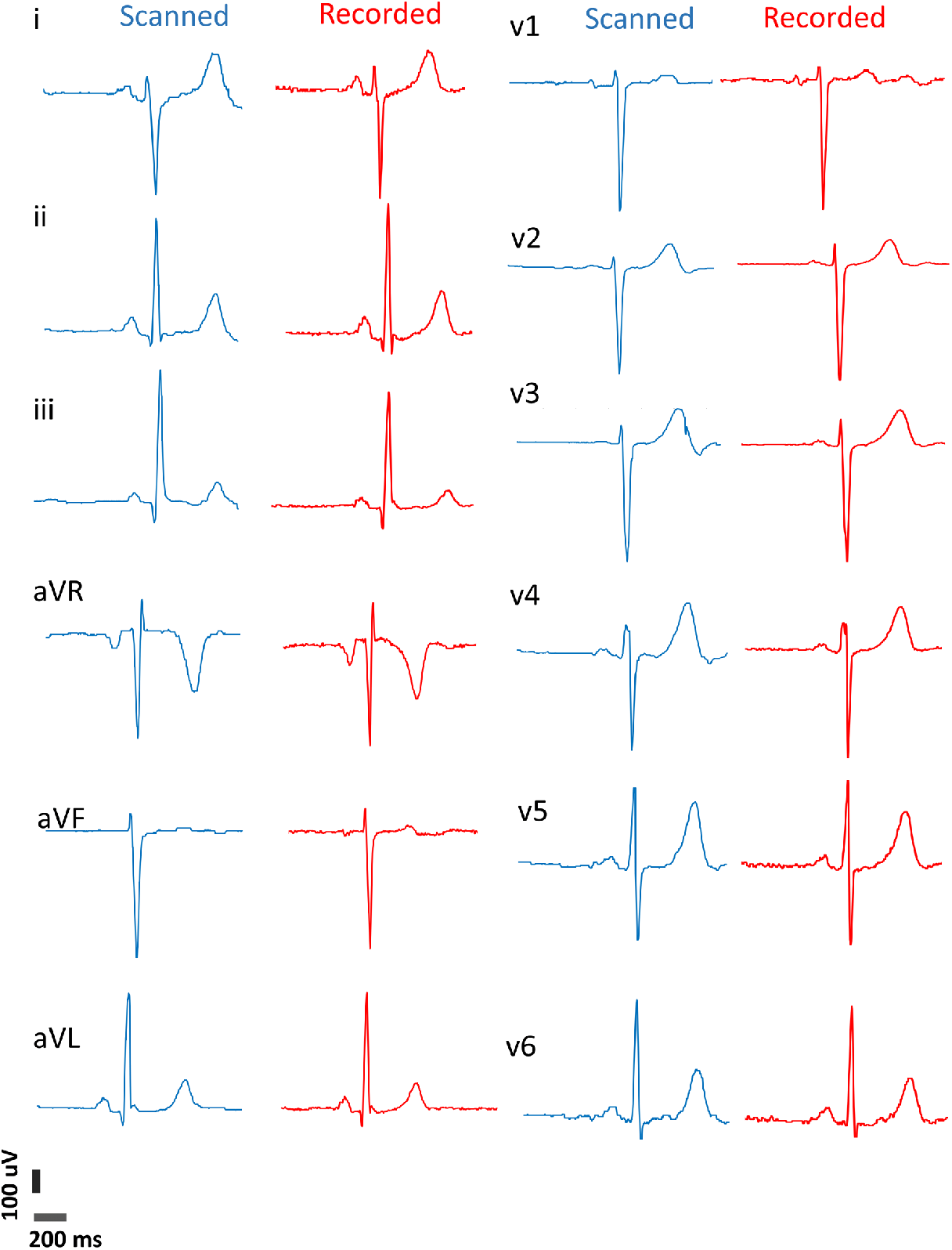
Comparison of a digitized (blue) and a digitally-recorded (red) 12-lead ECG signal.

#### Agreement between digitally recorded and digitized asynchronous representative beat

The precision of traditional ECG measurements (PR, QRS, QT intervals) assessed on digitized versus digitally recorded cardiac beat was substantial or nearly perfect, ranging from 97.2% to 99.1% (Table 3). Accordingly, the mean bias was small (up to 4 ms). However, their agreement measured by ρ_c_ was poor, and ICC was < 0.9, consistently with wide LOA (Table 3).

**Table 3.**
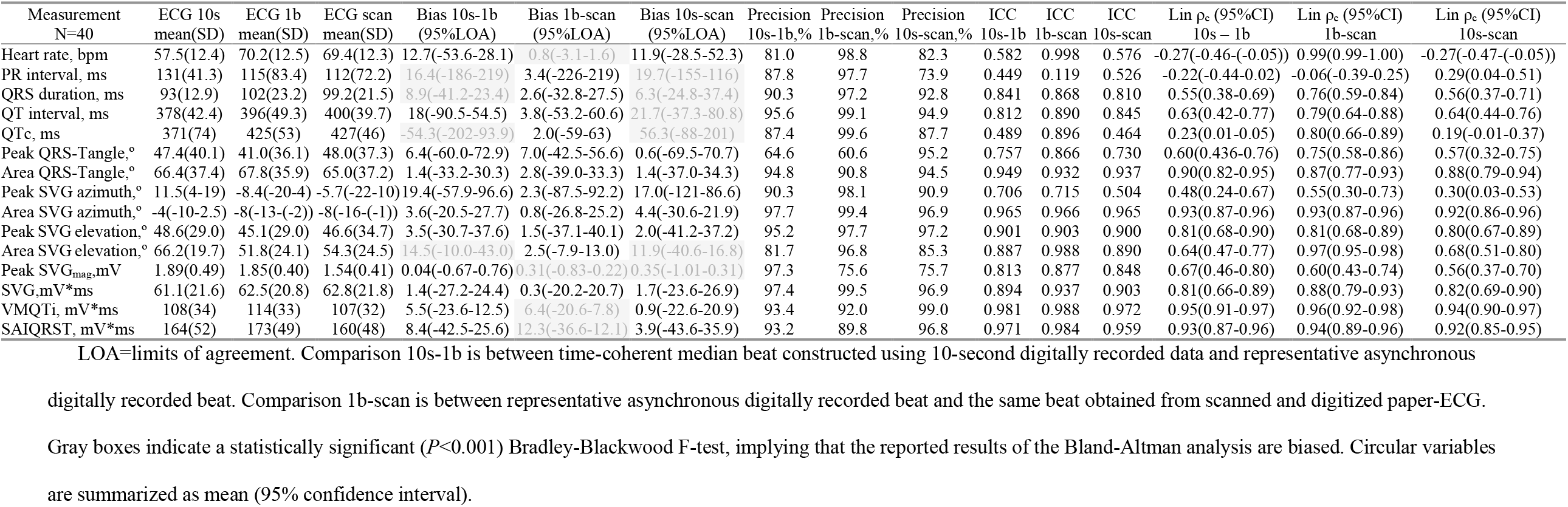
Agreement of ECG and VCG GEH measurements on digitally recorded and digitized ECG signal, in 3 types of beats.

The precision and agreement of area-based GEH VCG measurements were better than traditional ECG measurements, with ICC > 0.9 and ρ_c_ up to 0.97, thus indicating substantial agreement. Mean bias and 95%LOA were within a clinically acceptable range for area-based GEH metrics.

Notably, the precision and agreement of peak-based GEH VCG measurement were inadequate and unsatisfactory, with all coefficients below 0.9 (or less than 90%).

#### Agreement between digitally recorded asynchronous beat and time-coherent median beat

There was a substantial difference in agreement observed for area-based versus peak-based GEH VCG measurements. All but one area-based GEH VCG measurements showed substantial agreement, reflected by ICC and ρ_c_ >0.95 and high precision ≥ 95%. Mean bias for area-based VCG angles was minor (∼1°), with clinically acceptable 95%LOA (±20-30°). However, the mean bias for peak-based VCG angles was substantial (∼ 10°), and 95% LOA was clinically unacceptable (± 60-90°). One exception was SVG elevation, demonstrating higher precision and agreement if measured by peak-based (rather than area-based) metrics. (Table 3).

Precision and agreement for traditional ECG measurements ranged from poor to moderate.

#### Agreement between digitized asynchronous representative beat and digitally recorded time-coherent median beat

Out of all ECG and VCG measurements (Table 3), SAIQRST, VMQTi, and area-based SVG magnitude showed the highest agreement, precision, and concordance, the slightest mean bias (2-3 mV*ms), and clinically acceptable 95%LOA (±20-30 mV*ms).

Precision and agreement for other area-based GEH VCG measurements were also acceptable, ranging from moderate to substantial, with clinically acceptable bias. However, the agreement and precision for peak-based GEH VCG metrics were poor, and bias was unacceptably high.

A precision, agreement, and concordance observed for traditional ECG metrics were similar to those reported for pairs of digitally recorded asynchronous beat and time-coherent median beat.

## 4. Discussion

In this work, we developed and validated a new algorithm for paper-ECG digitization, converting a scan (i.e., image) of printed on paper ECG into a digital ECG signal. We implemented a novel dynamic programming approach with fully automated extraction of the digitized ECG signal, preserving its high resolution. For the first time, we provided open-source Python code, facilitating further development and implementation of the developed ECG digitization tool. Furthermore, we compared the sources of disagreement between ECG and VCG measurements performed on a digitally recorded and digitized ECG signal. We identified the morphological differences between median beats constructed using digitally, simultaneously recorded 8 ECG leads versus asynchronous ECG leads as the major source of the disagreement. Notably, in successfully digitized ECG signal, SAIQRST, VMQTi, and other area-based GEH VCG metrics showed substantial, clinically acceptable agreement. In contrast, peak-based VCG metrics as well as PR, QRS, and QT intervals demonstrated moderate to a poor agreement. As reported in the present study, bias with 95%LOA should be considered by future investigators planning analyses of digitized ECG.

### Comparison of the paper-ECG digitization methods

Several other algorithms for paper ECG digitization have been previously developed.[6, 7, 30-38] Unlike several earlier digitizing applications that scaled-down image resolution to reduce the computational cost,[7, 31] we elected to use high-resolution images (600 dpi), and output high-resolution ECG signal, taking advantage of computationally-efficient Python libraries.[18] Furthermore, we preserved the high resolution of the digitized ECG signal, providing users an opportunity to implement their preferred subsequent ECG signal processing approach.

Importantly, our algorithm did not “remove the grid from the ECG signal” but instead utilized a different approach – simultaneous extraction of both types of signals – grid and ECG waveform. Therefore, we did not encounter the removal of the ECG signal features observed by previous investigators.[31, 34]

Several recent algorithms incorporated somewhat similar approaches.[30, 35] However, our algorithm incorporated the most recent Python libraries, utilizing the advantages of the *Viterbi* dynamic programming.

Nonetheless, we acknowledge the shortcomings of the presented tool. We were able to successfully digitize only 50% of the ECGs in the validation dataset, whereas the digitization quality of another 50% of the data was deemed visually unacceptable. To provide the path forward and facilitate further development of paper-ECG digitizing tools, we provided our developed software code on GitHub, which is a convenient environment for software developers.

We acknowledge that the signal extraction algorithm does not perform well at sharp turning points (e.g., near the peaks of the QRS complex), which is a common problem in ECG digitization.[6, 7] In the future, this may be addressed by (1) using active contours with a dynamic low-pass filter for signal extraction or (2) adding a post-processing step that identifies turning points and pulls the estimated signal to the pixels at the outer edge of the turning point.

Furthermore, there may be errors in the signal extraction algorithm at the left and right edges. This may be addressed by (1) running *Viterbi* forward and backward, or (2) using active contours for signal extraction, or (3) implementing a more intelligent approach to find the best path.

### Simultaneously recorded versus asynchronous ECG leads

As expected, we observed significant differences in VCG morphology between a median beat comprised of 10-second digitally recorded simultaneous ECG leads and a representative cardiac beat constructed of digitally recorded, asynchronous ECG leads. Differences in VCG morphology translated into the differences in ECG and VCG measurements that have to be considered when comparing the results of different studies. Importantly, we observed clinically meaningful bias in QT interval measurements (∼ 20 ms). Similarly, Holkeri et al.[36] compared QT interval measured on asynchronous ECG leads and observed clinically important differences (>20 ms) in nearly 25% of their study population. Furthermore, Hingorani et al.[8] described additional contributions of the printer that affected differences in ECG measurements.

In the present study, we conducted detailed and the most comprehensive quantitative assessment of agreement between ECG and VCG measurements, performed on three different types of cardiac beat: (1) time-coherent median beat constructed using 10-second digitally recorded, simultaneous ECG leads, (2) digitally recorded, asynchronous ECG leads, and (3) digitized, asynchronous ECG leads. We reported mean bias with 95%LOA, precision, ICC, and concordance coefficients. As reported in this study, differences between different types of ECG signals have to be considered for planning future studies and interpreting previously conducted analyses. While further improvement in the digitization algorithms can reduce an error of the digitization procedure itself, fundamental differences between simultaneously recorded and asynchronous ECG leads reman. Area-based GEH VCG metrics appear to be robust and insensitive to different types of ECG signals and can be recommended for measurement on digitized ECGs. In contrast, peak-based VCG metrics are not recommended for measurement on digitized ECGs due to substantial disagreement between different types of ECG data.

### Limitations

Several limitations of the study have to be considered. First, while we attempted to comprise the diverse Development dataset, including both normal ECGs of healthy individuals, and abnormal ECGs of patients with cardiovascular diseases, printed on paper with variable degree of quality, there is a chance that our Development dataset did not include all possible types of ECG images. Therefore, different thresholds of the algorithm’s decisions might improve its performance on different types of paper-ECG images. Future studies are needed to address this limitation.

## 5. Conclusions

The newly developed and validated algorithm for paper-ECG digitization convert an image printed on paper ECG into a digital ECG signal with high precision. Differences in VCG morphology on time-coherent median beat comprised of 10-second simultaneously recorded ECG leads and a single cardiac beat comprised of asynchronous ECG leads contribute the most to the disagreement between ECG measurements on digitally recorded and digitized ECG. Area-based (but not peak-based) GEH VCG metrics preserve the clinically acceptable agreement and can be recommended for measurement on digitized ECG.

## Data Availability

The algorithm and open-source software code, written in Python (Python Software Foundation, Delaware, US), are provided at https://github.com/Tereshchenkolab/paper-ecg.

## Funding and Acknowledgment

This work was partially supported by the National Institutes of Health (HL118277), Medical Research Foundation of Oregon, and OHSU President Bridge funding to Tereshchenko.

## References

[1] L.G. Tereshchenko, N. Sotoodehnia, C.M. Sitlani, F.N. Ashar, M. Kabir, M.L. Biggs, M.P. Morley, J.W. Waks, E.Z. Soliman, A.E. Buxton, T. Biering-Sorensen, S.D. Solomon, W.S. Post, T.P. Cappola, D.S. Siscovick, D.E. Arking, Genome-Wide Associations of Global Electrical Heterogeneity ECG Phenotype: The ARIC (Atherosclerosis Risk in Communities) Study and CHS (Cardiovascular Health Study), J Am Heart Assoc, 7 (2018) e008160.

[2] J.D. Pollard, K.T. Haq, K.J. Lutz, N.M. Rogovoy, K.A. Paternostro, E.Z. Soliman, J. Maher, J.A.C. Lima, S.K. Musani, L.G. Tereshchenko, Electrocardiogram machine learning for detection of cardiovascular disease in African Americans: the Jackson Heart Study, Eur Heart J Digit Health, 2 (2021) 137–151.

[3] J.W. Waks, C.M. Sitlani, E.Z. Soliman, M. Kabir, E. Ghafoori, M.L. Biggs, C.A. Henrikson, N. Sotoodehnia, T. Biering-Sorensen, S.K. Agarwal, D.S. Siscovick, W.S. Post, S.D. Solomon, A.E. Buxton, M.E. Josephson, L.G. Tereshchenko, Global Electric Heterogeneity Risk Score for Prediction of Sudden Cardiac Death in the General Population: The Atherosclerosis Risk in Communities (ARIC) and Cardiovascular Health (CHS) Studies, Circulation, 133 (2016) 2222–2234.

[4] R. Brisk, R. Bond, E. Banks, A. Piadlo, D. Finlay, J. McLaughlin, D. McEneaney, Deep learning to automatically interpret images of the electrocardiogram: Do we need the raw samples?, Journal of Electrocardiology, 57 (2019) S65–S69.

[5] L.G. Tereshchenko, Global Electrical Heterogeneity: Mechanisms and Clinical Significance, Computing in Cardiology Conference (CinC), 45 (2018) e165

[6] G.S. Waits, E.Z. Soliman, Digitizing paper electrocardiograms: Status and challenges, J Electrocardiol, 50 (2017) 123–130.

[7] F. Badilini, T. Erdem, W. Zareba, A.J. Moss, ECGScan: a method for conversion of paper electrocardiographic printouts to digital electrocardiographic files, J Electrocardiol, 38 (2005) 310–318.

[8] P. Hingorani, D.R. Karnad, G.K. Panicker, S. Deshmukh, S. Kothari, D. Narula, Differences between QT and RR intervals in digital and digitized paper electrocardiograms: contribution of the printer, scanner, and digitization process, Journal of Electrocardiology, 41 (2008) 370–375.

[9] J.E. Norman, J.J. Bailey, A.S. Berson, W.K. Haisty, D. Levy, P.M. Macfarlane, P.M. Rautaharju, NHLBI workshop on the utilization of ECG databases: Preservation and use of existing ECG databases and development of future resources, Journal of Electrocardiology, 31 (1998) 83–89.

[10] P. Kligfield, L.S. Gettes, J.J. Bailey, R. Childers, B.J. Deal, E.W. Hancock, H.G. van, J.A. Kors, P. Macfarlane, D.M. Mirvis, O. Pahlm, P. Rautaharju, G.S. Wagner, M. Josephson, J.W. Mason, P. Okin, B. Surawicz, H. Wellens, Recommendations for the standardization and interpretation of the electrocardiogram: part I: the electrocardiogram and its technology a scientific statement from the American Heart Association Electrocardiography and Arrhythmias Committee, Council on Clinical Cardiology; the American College of Cardiology Foundation; and the Heart Rhythm Society endorsed by the International Society for Computerized Electrocardiology, J Am Coll.Cardiol., 49 (2007) 1109–1127.

[11] M.M. Kabir, E.A. Perez-Alday, J. Thomas, G. Sedaghat, L.G. Tereshchenko, Optimal configuration of adhesive ECG patches suitable for long-term monitoring of a vectorcardiogram, J Electrocardiol, 50 (2017) 342–348.

[12] J.A. Thomas, E.A. Perez-Alday, C. Hamilton, M.M. Kabir, E.A. Park, L.G. Tereshchenko, The utility of routine clinical 12-lead ECG in assessing eligibility for subcutaneous implantable cardioverter defibrillator, Comput Biol Med, 102 (2018) 242–250.

[13] J.A. Thomas, A.P.-A. E, A. Junell, K. Newton, C. Hamilton, Y. Li-Pershing, D. German, A. Bender, L.G. Tereshchenko, Vectorcardiogram in athletes: The Sun Valley Ski Study, Ann Noninvasive Electrocardiol, 24 (2019) e12614.

[14] L. Wang, N. Javadekar, A. Rajagopalan, N.M. Rogovoy, K.T. Haq, C.S. Broberg, L.G. Tereshchenko, Eligibility for subcutaneous implantable cardioverter-defibrillator in congenital heart disease, Heart Rhythm, 17 (2020) 860–869.

[15] PyQt5 is copyright (c) Riverbank Computing Limited. https://pypi.org/project/PyQt5/. Accessed 06.24.2021.

[16] G. Bradski, OpenCV. Open Source Computer Vision Library, Dr. Dobb’s Journal of Software Tools., (2000).

[17] Python: cv2.adaptiveThreshold. https://docs.opencv.org/2.4/modules/imgproc/doc/miscellaneous_transformations.html. Accessed 05.17.2021.

[18] P. Virtanen, R. Gommers, T.E. Oliphant, M. Haberland, T. Reddy, D. Cournapeau, E. Burovski, P. Peterson, W. Weckesser, J. Bright, S.J. van der Walt, M. Brett, J. Wilson, K.J. Millman, N. Mayorov, A.R.J. Nelson, E. Jones, R. Kern, E. Larson, C.J. Carey, I. Polat, Y. Feng, E.W. Moore, J. VanderPlas, D. Laxalde, J. Perktold, R. Cimrman, I. Henriksen, E.A. Quintero, C.R. Harris, A.M. Archibald, A.H. Ribeiro, F. Pedregosa, P. van Mulbregt, C. SciPy, SciPy 1.0: fundamental algorithms for scientific computing in Python, Nature methods, 17 (2020) 261–272.

[19] N. Otsu, A Threshold Selection Method from Gray-Level Histograms, IEEE Transactions on Systems, Man, and Cybernetics, 9 (1979) 62–66.

[20] L. Huang, Advanced Dynamic Programming in Semiring and Hypergraph Frameworks, Coling 2008 Organizing Committee, Manchester, UK, 2008, pp. 1–18.

[21] E.A. Perez-Alday, Y. Li-Pershing, A. Bender, C. Hamilton, J.A. Thomas, K. Johnson, T.L. Lee, R. Gonzales, A. Li, K. Newton, L.G. Tereshchenko, Importance of the heart vector origin point definition for an ECG analysis: The Atherosclerosis Risk in Communities (ARIC) study, Comput Biol Med, 104 (2019) 127–138.

[22] J. Pan, W. Tompkins, A real-time QRS detection algorithm, IEEE transactions on bio-medical engineering, 32 (1985) 230–236.

[23] W. Zong, M. Saeed, T. Heldt, A QT interval detection algorithm based on ECG curve length transform, Comput.Cardiol., 33 (2006) 377–380.

[24] E.A. Perez-Alday, A. Bender, D. German, S.V. Mukundan, C. Hamilton, J.A. Thomas, Y. Li-Pershing, L.G. Tereshchenko, Dynamic predictive accuracy of electrocardiographic biomarkers of sudden cardiac death within a survival framework: the Atherosclerosis Risk in Communities (ARIC) study, BMC cardiovascular disorders, 19 (2019) 255.

[25] J.A. Kors, H.G. van, A.C. Sittig, J.H. van Bemmel, Reconstruction of the Frank vectorcardiogram from standard electrocardiographic leads: diagnostic comparison of different methods, Eur.Heart J, 11 (1990) 1083–1092.

[26] J.M. Bland, D.G. Altman, Statistical methods for assessing agreement between two methods of clinical measurement, Lancet, 1 (1986) 307–310.

[27] T. Huang, C.A. James, C. Tichnell, B. Murray, J. Xue, H. Calkins, L.G. Tereshchenko, Statistical evaluation of reproducibility of automated ECG measurements: an example from arrhythmogenic right ventricular dysplasia/cardiomyopathy clinic, Biomedical signal processing and control, 13 (2014) 23–30.

[28] E.L. Bradley, L.G. Blackwood, Comparing Paired Data: A Simultaneous Test for Means and Variances, The American Statistician, 43 (1989) 234–235.

[29] G. Bravo, L. Potvin, Estimating the reliability of continuous measures with Cronbach’s alpha or the intraclass correlation coefficient: toward the integration of two traditions, Journal of clinical epidemiology, 44 (1991) 381–390.

[30] Y. Li, Q. Qu, M. Wang, L. Yu, J. Wang, L. Shen, K. He, Deep learning for digitizing highly noisy paper-based ECG records, Computers in Biology and Medicine, 127 (2020) 104077.

[31] L. Ravichandran, C. Harless, A.J. Shah, C.A. Wick, J.H. Mcclellan, S. Tridandapani, Novel tool for complete digitization of paper electrocardiography data, IEEE journal of translational engineering in health and medicine, 1 (2013) 1800107–1800107.

[32] A. Sbrollini, A. Agostinelli, I. Marcantoni, M. Morettini, L. Burattini, F. Di Nardo, S. Fioretti, L. Burattini, eCTG: an automatic procedure to extract digital cardiotocographic signals from digital images, Computer methods and programs in biomedicine, 156 (2018) 133–139.

[33] S. Wang, S. Zhang, Z. Li, L. Huang, Z. Wei, Automatic digital ECG signal extraction and normal QRS recognition from real scene ECG images, Computer methods and programs in biomedicine, 187 (2020) 105254.

[34] P. Swamy, S. Jayaraman, M.G. Chandra, An improved method for digital time series signal generation from scanned ECG records, 2010 International Conference on Bioinformatics and Biomedical Technology, 2010, pp. 400–403.

[35] M. Baydoun, L. Safatly, O.K.A. Hassan, H. Ghaziri, A.E. Hajj, H. Isma’eel, High Precision Digitization of Paper-Based ECG Records: A Step Toward Machine Learning, IEEE Journal of Translational Engineering in Health and Medicine, 7 (2019) 1–8.

[36] A. Holkeri, A. Eranti, T.V. Kenttä, K. Noponen, M.A.E. Haukilahti, T. Seppänen, M.J. Junttila, T. Kerola, H. Rissanen, M. Heliövaara, P. Knekt, A.L. Aro, H.V. Huikuri, Experiences in digitizing and digitally measuring a paper-based ECG archive, Journal of Electrocardiology, 51 (2018) 74–81.

[37] T. Kao, H. Len-Jon, L. Yui-Han, L. Tzong-Huei, H. Chia-Hung, Computer analysis of the electrocardiograms from ECG paper recordings, 2001 Conference Proceedings of the 23rd Annual International Conference of the IEEE Engineering in Medicine and Biology Society, 2001, pp. 3232-3234 vol.3234.

[38] J.T. Wang, D.P. Mital, A microcomputer-based prototype for ECG paper record conversion, Journal of Network and Computer Applications, 19 (1996) 295–307.

